# Prognostic significance of baseline low-density lipoprotein cholesterol in patients undergoing coronary revascularization; A report from the CREDO-Kyoto registry

**DOI:** 10.1101/2023.10.26.23297646

**Authors:** Kenji Kanenawa, Kyohei Yamaji, Takeshi Morimoto, Ko Yamamoto, Takenori Domei, Makoto Hyodo, Hiroki Shiomi, Yutaka Furukawa, Yoshihisa Nakagawa, Kazushige Kadota, Hirotoshi Watanabe, Yusuke Yoshikawa, Tomohisa Tada, Junichi Tazaki, Natsuhiko Ehara, Ryoji Taniguchi, Toshihiro Tamura, Atsushi Iwakura, Takeshi Tada, Satoru Suwa, Mamoru Toyofuku, Tsukasa Inada, Kazuhisa Kaneda, Tatsuya Ogawa, Teruki Takeda, Hiroshi Sakai, Takashi Yamamoto, Keiichi Tambara, Jiro Esaki, Hiroshi Eizawa, Miho Yamada, Eiji Shinoda, Junichiro Nishizawa, Hiroshi Mabuchi, Nobushige Tamura, Manabu Shirotani, Shogo Nakayama, Takashi Uegaito, Mitsuo Matsuda, Mamoru Takahashi, Moriaki Inoko, Naoki Kanemitsu, Takashi Tamura, Kazuhisa Ishii, Ryuzo Nawada, Tomoya Onodera, Nobuhisa Ohno, Tadaaki Koyama, Hiroshi Tsuneyoshi, Hiroki Sakamoto, Takeshi Aoyama, Shinji Miki, Masaru Tanaka, Yukihito Sato, Fumio Yamazaki, Michiya Hanyu, Yoshiharu Soga, Tatsuhiko Komiya, Kenji Minatoya, Kenji Ando, Takeshi Kimura

## Abstract

**Background:** The impact of very low baseline levels of low-density lipoprotein cholesterol (LDL-C) on patients with coronary artery disease remains unclear. Therefore, we aimed to investigate the baseline characteristics and clinical outcomes of patients with low baseline LDL-C levels who had undergone coronary revascularization.

**Methods:** We enrolled 39439 patients of the pooled population from the CREDO-Kyoto registries Cohorts 1, 2, and 3. After excluding 6306 patients with missing baseline LDL-C data, the study population consisted of 33133 patients who had undergone their first coronary revascularization. We assessed the risk for mortality and cardiovascular events according to quintiles of the baseline LDL-C levels.

**Results:** Patients in the very low LDL-C quintile (< 85 mg/dL) had more comorbidities than those in the other quintiles. Lower LDL-C levels were strongly associated with anemia, thrombocytopenia, and end-stage renal disease. The cumulative 4-year incidence of all-cause death increased as LDL-C levels decreased (very low: 19.4%, low: 14.5%, intermediate: 11.1%, high: 10.0%, and very high:9.2%; P<0.001), which was driven by both the early and late events. After adjusting for baseline characteristics, the adjusted risks of the very low and low LDL-C quintiles relative to the intermediate LDL-C quintile remained significant for all-cause death (very low: HR 1.29, 95% CI 1.16-1.44, P<0.001; low: HR 1.15, 95% CI 1.03-1.29, P=0.01). There were no significant interactions between the association of LDL-C level with all-cause death and subgroup factors, such as lipid-lowering treatment at index hospitalization, age, sex, acute myocardial infarction presentation, and study cohort. The excess adjusted risks of the lowest LDL-C quintile relative to the intermediate LDL-C quintile were significant for clinical outcomes such as cardiovascular death (HR 1.17, 95% CI 1.01-1.35), non-cardiovascular death (HR 1.35, 95% CI 1.15-1.60), sudden death (HR 1.44, 95% CI 1.01-2.06), and heart failure admission (HR 1.11 95% CI 1.01-1.22), while there was no excess risk for the lowest LDL-C quintile relative to the intermediate LDL-C quintile for myocardial infarction and stroke.

**Conclusions:** Lower baseline LDL-C levels were associated with more comorbidities and a significantly higher risk of death, regardless of cardiovascular or non-cardiovascular causes, in patients who underwent coronary revascularization.

## CLINICAL PERSPECTIVE

### What is new?

- This is the first study to show that among patients undergoing coronary revascularization, those with very low baseline LDL-C levels have more comorbidities and poor clinical outcomes in those who underwent revascularization.
- These poor outcomes include cardiovascular, non-cardiovascular, sudden death and heart failure admission.

### What are the clinical implications?

- Patients with coronary artery disease and low LDL-C levels need to be managed with special care.
- Special attention should be given not only to cardiovascular problems but also to non-cardiovascular problems.
- This is because these patients are more likely to experience both cardiovascular and non-cardiovascular events.

## Introduction

Lipid-lowering therapy for low-density lipoprotein cholesterol (LDL-C) is widely recommended to mitigate the risk of cardiovascular events,^1–3^ particularly among high-risk patients with a history of atherosclerotic cardiovascular disease (ASCVD).^4–8^ High-intensity statin therapy is considered the primary treatment of choice for these patients regardless of their baseline LDL-C levels. Moreover, several randomized controlled trials have demonstrated that additional LDL-C-lowering therapy with ezetimibe and/or proprotein convertase subtilisin-kexin type 9 (PCSK9) inhibitors further reduced the coronary artery plaque burden^9–11^ and provided further clinical benefit.^12–14^ Based on these studies, additional LDL-C-lowering therapy in addition to high-intensity statin therapy targeting an LDL-C level below 70 mg/dL is recommended in patients with clinical ASCVD who are judged to be very high risk. However, since achieved LDL-C levels were strongly correlated with baseline LDL-C levels, patients with lower baseline LDL-C levels often had LDL-C levels below 70 mg/dL after high-intensity statin therapy, and were less likely to be treated with additional LDL-C lowering therapy.^15^ Moreover, both low and high LDL-C levels in the general population were associated with an increased risk for all-cause death, with the lowest risk found at an LDL-C level of 140 mg/dL.^16^ However, it remained largely unclear whether patients with low baseline LDL-C levels who have a history of ASCVD and could be candidates for aggressive LDL-C-lowering therapy are also at a higher risk for all-cause death.^17^

Therefore, we aimed to investigate the baseline characteristics and clinical outcomes of patients with low baseline LDL-C levels who underwent coronary revascularization.

## Methods

### Study population

The Coronary Revascularization Demonstrating Outcome Study in Kyoto (CREDO-Kyoto) percutaneous coronary intervention (PCI) and coronary artery bypass grafting (CABG) registries are multicenter, physician-initiated, non-company-sponsored registries that enrolled consecutive patients who underwent their first coronary revascularization with PCI or isolated CABG. CREDO-Kyoto registry Cohort 1 enrolled 9,877 patients with coronary artery disease, excluding those with acute myocardial infarction (AMI), from 21 centers between January 1, 2000, and December 31, 2002, during the bare-metal stent era. Cohort 2 enrolled 15,939 patients, including those with AMI, from 26 centers between January 1, 2005, and December 31, 2007, after the introduction of drug-eluting stents in 2004. Cohort 3 enrolled 14,927 patients, including those with AMI, from 22 centers between January 1, 2011, and December 31, 2013, after the approval of new-generation drug-eluting stents in 2010. The analytic populations of the CREDO-Kyoto registries included 9341 patients in Cohort 1, 15231 patients in Cohort 2, and 14867 patients in Cohort 3, after excluding patients who had undergone combined noncoronary surgery, patients who refused to participate in the registry, and patients in Cohort 1 who presented with AMI (a violation of the inclusion criteria). We pooled the 3 cohorts with a total of 39439 patients. We further excluded 2578 patients in Cohort 1, 2946 patients in Cohort 2, and 782 patients in Cohort 3. The final study population comprised 33,133 patients (**Figure 1**).

**Figure 1.**
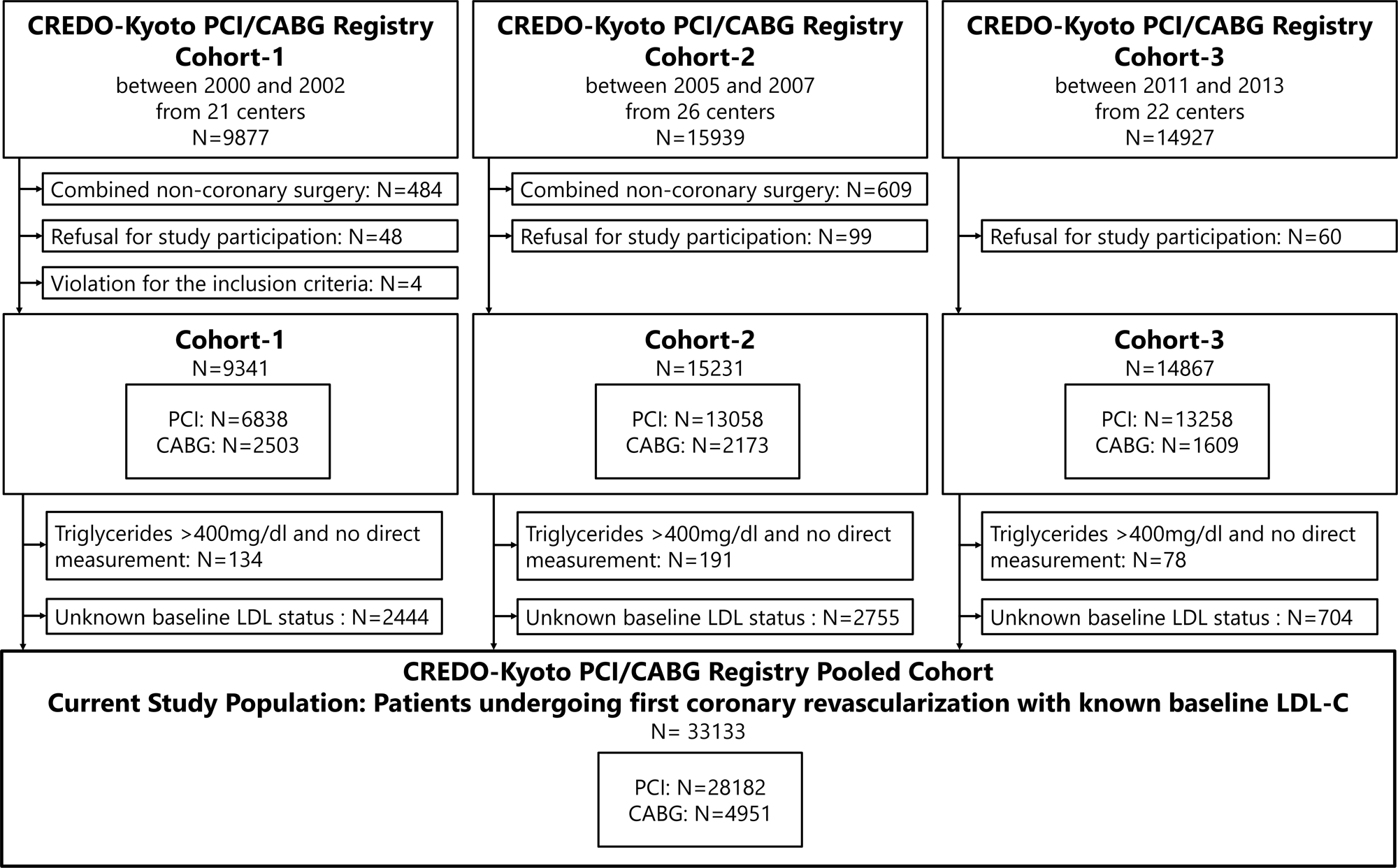
Study patient flow. CREDO-Kyoto=Coronary Revascularization Demonstrating Outcome Study in Kyoto, LDL-C=low-density lipoprotein cholesterol, PCI=percutaneous coronary intervention, CABG=coronary artery bypass grafting.

The relevant ethics committees of all the participating centers approved the study protocol. As enrollment was retrospective, the requirement for written informed consent was waived; however, we excluded the 207 patients who refused to participate in the study when contacted for follow-up. This approach was concordant with the guidelines of the Japanese Ministry of Health, Labor, and Welfare.

### Data collection for baseline characteristics and follow-up events

Clinical, angiographic, and procedural data were collected from the hospital charts or databases according to the pre-specified definitions by experienced clinical research coordinators from the Research Institute for Production Development (Kyoto, Japan) (**<u>Supplemental Appendix A</u>**). Blood examinations were performed before index PCI. Since lipid data are occasionally not available before the index PCI in emergency conditions such as AMI and cardiogenic shock, the use of lipid data during the hospitalization period was allowed in these registries. Information regarding lipid-lowering treatments was obtained separately at admission and discharge. LDL-C levels were measured directly or calculated according to the Friedewald equation if LDL-C levels were not measured and triglyceride concentrations were less than 400 mg/dL.

Follow-up data were collected from hospital charts or obtained through contact with the patients, their relatives, or the referring physicians. Clinical events after the index procedure were assessed as follow-up events, except for the scheduled staged coronary revascularization procedures performed within 3 months of the index procedure, which were regarded as part of the index procedure. The clinical event committee determined whether any incident was a clinical event (**<u>Supplemental Appendix B</u>**).

### Clinical outcome measures

We assessed all-cause death, cardiovascular death, non-cardiovascular death, myocardial infarction (MI), stroke, and admission for heart failure at 4 years. Definitions for the outcome measures were consistent across the three cohorts. Death was regarded as cardiac in origin unless obvious non-cardiac causes could be identified; thus, death from an unknown cause and any death during the index hospitalization for coronary revascularization were regarded as cardiac death. Cardiovascular death included cardiac and other deaths related to stroke, renal disease, and vascular diseases. MI was adjudicated according to the ARTS (Arterial Revascularization Therapies Study) definition, in which only Q-wave MI was regarded as MI when it occurred within 7 days of the index procedure.^18^ Stroke was defined as an ischemic or hemorrhagic stroke with neurological symptoms lasting longer than 24 hours. Admission for heart failure was defined as hospitalization for worsening heart failure requiring intravenous drug therapy.

### Statistical analysis

Continuous variables were expressed as mean ± standard deviation or as median (interquartile range) and compared using Student’s t-test or the Wilcoxon rank-sum test. Categorical variables were presented as values and percentages and were compared using the χ^2^ test. To assess trends in binary and continuous outcome variables, we utilized the Cochran–Armitage and Jonckheere–Terpstra trend tests, respectively. Kaplan–Meier curves were used to estimate the cumulative incidence of clinical events, and patients who had no clinical events at 4 years were censored. To assess the effect of LDL-C levels on clinical outcomes, we stratified patients into quintiles based on their LDL-C levels. The hazard ratios (HRs) and 95% confidence intervals (CIs) for patients with various LDL-C levels were estimated using univariate and multivariable Cox regression models. To adjust for confounders, we included the following explanatory variables into the multivariable models: age, sex, body mass index (BMI), AMI presentation, LDL-C lowering treatment at discharge, smoking status, hypertension, diabetes, acute heart failure, cardiogenic shock, ejection fraction, prior peripheral artery disease, prior heart failure, prior stroke, atrial fibrillation, chronic obstruction pulmonary disease, liver cirrhosis, malignancy, estimated glomerular filtration rate (eGFR), hemoglobin, platelet count, cohort number, use of β blocker, use of renin-angiotensin system inhibitors, and use of any medications for dyslipidemia at hospitalization.

To minimize the effects of early events related to PCI on clinical outcomes, we conducted 3-month landmark analyses. Additionally, we conducted subgroup analyses to investigate potential interactions among different patient subgroups, such as lipid-lowering treatment, age (≥75 years), sex, AMI presentation, and study cohorts. To estimate the interaction effects among these subgroups, we calculated the Type III sum of squares. For the sensitivity analysis. We divided the patients into seven groups according to the percentiles of LDL-C distribution (5^th^, 20^th^, 40^th^, 60^th^, 80^th^, and 95^th^ percentiles). To visualize the potential nonlinear relationships between LDL-C levels and clinical outcomes, we constructed multivariable Cox regression models with penalized smoothing splines and degrees of freedom determined using the lowest Akaike information criterion.^19, 20^

We constructed a gradient boosting model to estimate the risk factors with the strongest association with LDL-C according to their relative importance (RI). The gradient boosting model, a machine learning method, uses decision trees as weak learners, and bagging and boosting techniques are used taking into account the complex higher-order interactions to provide robustness, bias reduction, and higher prediction accuracy. Furthermore, a two-way partial dependence plot (PDP) based on the gradient boosting model, which showed the marginal effect of the two predictors on the prediction, was used to visualize the association and interaction between LDL-C and the other predictors of all-cause death.^21–23^ PDP simplifies input-output relationships by marginalizing and substantially eliminating the effects of variables other than those of interest.

Risk factors other than LDL-C in the PDP were selected based on their impact on mortality and the requirement for them to be continuous values.

All analyses were performed using R software (version 4.1.1; R Foundation for Statistical Computing, Vienna, Austria) and JMP version 14.3.0 (SAS Institute Incorporated, Cary, NC, USA).

## Results

The mean age was 68.3±10.8 years, and men accounted for 72.3% of the study population. The median (quartiles) of LDL-C level was 112 (90.2, 135) mg/dL. Patients were divided into quintiles based on the LDL-C level (Lowest group [LDL-C <85 mg/dL]: N=6599, Lower group [85 mg/dL≤ LDL-C <105 mg/dL]: N=6818, Intermediate group [105 mg/dL≤ LDL-C <120 mg/dL]: N=6299, Higher group [120 mg/dL≤ LDL-C <141 mg/dL]: N=6747, and the Highest group [141 mg/dL≤ LDL-C]: N=6747). The baseline clinical characteristics according to the LDL-C quintiles are summarized in **Table 1**. Compared with patients in the other groups, those in the lowest LDL-C group were older, more often men, had a lower BMI, and more often presented with cardiogenic shock. Patients in the lowest LDL-C group more often had other comorbidities, including renal failure, dialysis, chronic obstructive pulmonary disease, prior heart failure, lower ejection fraction, atrial fibrillation, liver cirrhosis, malignancy, anemia, and thrombocytopenia than those in the other LDL-C groups. The rate of lipid-lowering therapy use was 38.6% at admission and 71.5% at discharge.

**Table 1.** Baseline Characteristics in the 5 groups according to the baseline LDL-C levels.

|  | Overall<br>(N=33133) | Baseline LDL-C (mg/dl) |  |  |  |  | P values | P for trend |
| --- | --- | --- | --- | --- | --- | --- | --- | --- |
|  |  | Quintiles |  |  |  |  |  |  |
|  |  | Lowest<br>group<br>(LDL-C <85) | Lower<br>group<br>(85≤ LDL-C <105) | Intermediate<br>group<br>(105≤ LDL-C <120) | Higher<br>group<br>(120≤ LDL-C <141) | Highest<br>group<br>(141≤ LDL-C) |  |  |
|  |  | N=6599 | N=6818 | N=6299 | N=6747 | N=6670 |  |  |
| Patient characteristics |  |  |  |  |  |  |  |  |
| Age, year | 68.3±10.8 | 69.9±10.8 | 69.7±10.4 | 68.7±10.5 | 68.0±10.6 | 65.9±11.0 | <0.001 | <0.001 |
| Men | 22475 (72.3%) | 5050 (76.5%) | 5007 (73.4%) | 4665 (74.1%) | 4840 (71.7%) | 4420 (66.3%) | <0.001 | <0.001 |
| Body mass index | 23.8±3.5 | 23.4±3.5 | 23.5±3.5 | 23.7±3.4 | 24.0±3.5 | 24.2±3.4 | <0.001 | <0.001 |
| Acute myocardial infarction | 9116 (27.5%) | 1740 (26.4%) | 1780 (26.1%) | 1662 (26.4%) | 1868 (27.7%) | 2065 (31.0%) | <0.001 | <0.001 |
| ST-segment elevation myocardial infarction | 7294 (22.0%) | 1394 (21.1%) | 1444 (21.2%) | 1314 (20.9%) | 1493 (22.1%) | 1649 (24.7%)) | <0.001 | <0.001 |
| Non-ST-segment elevation myocardial infarction | 1822 (5.5%) | 346 (5.3%) | 336 (4.9%) | 348 (5.5%) | 375 (5.6%) | 416 (6.3%) | <0.001 | <0.001 |
| Cardiogenic shock | 580 (1.8%) | 191 (2.9%) | 128 (1.9%) | 80 (1.3%) | 89 (1.3%) | 92 (1.4%) | <0.001 | <0.001 |
| Prior stroke | 4127 (12.5%) | 968 (14.7%) | 953 (14.0%) | 792 (12.6%) | 764 (11.3%) | 650 (9.8%) | <0.001 | <0.001 |
| Hypertension | 26494 (80.0%) | 5450 (82.6%) | 5545 (81.3%) | 5027 (79.8%) | 5381 (80.0%) | 5091 (76.3%) | <0.001 | <0.001 |
| Diabetes mellitus | 13001 (39.3%) | 3115 (47.2%) | 2771 (40.7%) | 2446 (38.8%) | 2414 (35.8%) | 2255 (33.8%) | <0.001 | <0.001 |
| on insulin therapy | 2905 (8.8%) | 900 (13.7%) | 649 (9.5%) | 521 (8.3%) | 432 (6.4%) | 406 (6.1%) | <0.001 | <0.001 |
| Current smoker | 9405 (28.5%) | 1677 (25.5%) | 1806 (26.6%) | 1816 (28.9%) | 1939 (28.9%) | 2167 (32.6%) | <0.001 | <0.001 |
| Renal failure |  |  |  |  |  |  |  |  |
| eGFR <30 mL/min per 1.73m<sup>2</sup> without dialysis | 1448 (4.4%) | 402 (6.1%) | 326 (4.8%) | 244 (3.9%) | 228 (3.4%) | 248 (3.7%) | <0.001 | <0.001 |
| Dialysis | 1354 (4.1%) | 579 (8.8%) | 296 (4.3%) | 195 (3.1%) | 173 (2.6%) | 111 (1.7%) | <0.001 | <0.001 |
| Chronic obstructive pulmonary disease | 1159 (3.5%) | 264 (4.0%) | 243 (3.6%) | 230 (3.7%) | 204 (3.0%) | 218 (3.3%) | 0.03 | 0.03 |
| Peripheral artery disease | 3034 (9.2%) | 767 (11.6%) | 669 (9.8%) | 611 (9.7%) | 531 (7.9%) | 456 (6.8%) | <0.001 | <0.001 |
| Prior heart failure | 1826 (5.5%) | 536 (8.1%) | 380 (5.6%) | 334 (5.3%) | 302 (4.5%) | 274 (4.1%) | <0.001 | <0.001 |
| Ejection fraction <40% | 2823 (8.5%) | 662 (10.0%) | 565 (8.3%) | 516 (8.2%) | 501 (7.4%) | 579 (8.7%) | <0.001 | <0.001 |
| Atrial fibrillation | 2943 (8.9%) | 750 (11.4%) | 679 (10.0%) | 561 (8.9%) | 528 (7.8%) | 425 (6.4%) | <0.001 | <0.001 |
| Liver cirrhosis | 883 (2.7%) | 727 (4.1%) | 208 (3.1%) | 162 (2.6%) | 132 (2.0%) | 109 (1.6%) | <0.001 | <0.001 |
| Malignancy | 3258 (9.8%) | 875 (13.3%) | 755 (11.1%) | 613 (9.7%) | 540 (8.0%) | 475 (7.1%) | <0.001 | <0.001 |
| Hemoglobin <11g/dl | 4101 (12.4%) | 1388 (21.0%) | 1003 (14.7%) | 652 (10.4%) | 593 (8.8%) | 465 (7.0%) | <0.001 | <0.001 |
| Thrombocytopenia (Platelet<100×10<sup>9</sup>/L) | 476 (1.4%) | 208 (3.2%) | 128 (1.9%) | 61 (1.0%) | 52 (0.8%) | 27 (0.4%) | <0.001 | <0.001 |
| Total cholesterol | 188.8±41.5 | 140.6±24.2 | 167.0±18.4 | 184.9±17.2 | 203.8±17.7 | 242.3±32.2 | <0.001 | <0.001 |
| High density lipoprotein | 47.9±13.7 | 47.0±15.6 | 47.7±13.8 | 48.0±13.0 | 48.3±12.9 | 48.7±12.5 | <0.001 | <0.001 |
| Triglyceride | 130.9±85.6 | 123.1±91.3 | 121.5±73.1 | 128.3±72.5 | 131.7±73.3 | 137.8±79.8 | <0.001 | <0.001 |
| **Procedural characteristics** |  |  |  |  |  |  |  |  |
| Multivessel coronary artery disease | 20204 (61.0%) | 4180 (63.3%) | 4147 (60.8%) | 3787 (60.1%) | 4006 (59.4%) | 4084 (61.2%) | <0.001 | <0.001 |
| Left main coronary artery disease | 2783 (8.4%) | 657 (10.0%) | 596 (8.7%) | 503 (8.0%) | 494 (7.3%) | 533 (8.0%) | <0.001 | <0.001 |
| **Medication at index hospitalization** |  |  |  |  |  |  |  |  |
| β blocker | 10709 (32.3%) | 2300 (34.9%) | 2184 (32.1%) | 1988 (31.6%) | 2127 (31.5%) | 2110 (31.7%) | <0.001 | <0.001 |
| Renin-angiotensin system inhibitors | 17707 (53.5%) | 3552 (53.8%) | 3695 (54.2%) | 3392 (53.9%) | 3562 (52.8%) | 3506 (52.3%) | 0.22 | 0.04 |
| Oral anticoagulant | 4292 (13.0%) | 1052 (15.9%) | 935 (13.7%) | 795 (12.6%) | 783 (11.6%) | 727 (10.9%) | <0.001 | <0.001 |
| Any medications for dyslipidemia | 12802 (38.6%) | 3271 (49.6%) | 2800 (41.1%) | 2243 (35.6%) | 2241 (33.2%) | 2247 (33.7%) | <0.001 | <0.001 |
| Statins | 11933 (36.0%) | 3168 (48.0%) | 2613 (38.3%) | 2071 (32.9%) | 2052 (30.4%) | 2029 (30.4%) | <0.001 | <0.001 |
| **Medication at discharge** |  |  |  |  |  |  |  |  |
| Any medications for dyslipidemia | 23701 (71.5%) | 4765 (72.2%) | 4671 (68.5%) | 4326 (68.7%) | 4727 (70.1%) | 5212 (78.1%) | <0.001 | <0.001 |
| Statins | 22439 (67.7%) | 4314 (65.3%) | 4415 (64.8%) | 4120 (65.4%) | 4541 (67.3%) | 5049 (75.7%) | <0.001 | <0.001 |

1 Values were missing for BMI in 522 patients, for diabetes in 6 patients, for hypertension in 3 patients, for smoker status in 122 patients, for eGFR in 243 patients, complete blood count in 4728 patients, for ejection fraction in 4484 patients, 2 and for beta blockers in 16 patients.
3 Continuous variables are given as mean ± standard deviation or median (interquartile range). Categorical variables are given as number (%).
4 LDL-C=low-density lipoprotein cholesterol, eGFR=estimated glomerular filtration rate.

Strong relationships were identified between low LDL-C levels and anemia, thrombocytopenia, and dialysis. To further clarify the relationship, each strong predictor was evaluated according to its relative importance calculated using a gradient boosting model (**Supplemental Figure 1**). In this model, a low LDL-C level was defined as 105 mg/dl or less. While the three strongest predictors of low LDL-C levels were anemia, thrombocytopenia, and dialysis, a low LDL-C level was not a strong predictor of anemia, dialysis, or low platelet count.

### Clinical outcomes

Clinical follow-up information at 4 years was obtained for 30937 (93.4%) patients. The cumulative 4-year incidence of clinical outcomes is presented in **Table 2**. The cumulative 4-year incidence of all-cause death increased with decreasing LDL-C levels (very low,19.4%; low,14.5%; intermediate,11.1%; high, 10.0%; and very high,9.2%; P < 0.001), as did the incidences of cardiovascular death, non-cardiovascular death, stroke, and admission for heart failure (**Figure 2**). In the 3-month landmark analyses, an increasing incidence of all-cause death with decreasing LDL-C levels were consistently seen both within and beyond 3 months. In the univariate Cox regression analysis, the crude risk for all-cause death incrementally increased from the highest LDL-C group (HR 0.82, 95% CI 0.74-0.92, P<0.001) to the lowest LDL-C group (HR 1.86, 95% CI 1.70-2.05, P<0.001) with the intermediate LDL-C group as the reference (**Table 3 and Figure 3**). After adjusting for baseline characteristics, the excess risks of the lowest and lower LDL-C groups relative to the intermediate LDL-C group remained significant for all-cause death (lowest: adjusted HR 1.29, 95% CI 1.16-1.44, P<0.001; lower: adjusted HR 1.15, 95% CI 1.03-1.29, P=0.01), while the excess risks of the higher and highest LDL-C groups relative to the intermediate LDL-C group were not significant for all-cause death (higher: adjusted HR 1.00, 95% CI 0.93-1.19, P=0.45; highest: adjusted HR 1.05, 95% CI 0.97-1.25, P=0.15). The excess adjusted risks of the lowest LDL-C group relative to the intermediate LDL-C group were also significant for clinical outcomes such as cardiovascular death (adjusted HR 1.17, 95% CI 1.01-1.35), non-cardiovascular death (adjusted HR 1.35, 95% CI 1.15-1.60), sudden death (adjusted HR 1.44, 95% CI 1.01-2.06), and admission for heart failure (adjusted HR 1.11 95% CI 1.01-1.22), while there was no excess risk of the lowest LDL-C group relative to the intermediate LDL-C group for MI and stroke (**Table 3**).

**Figure 2.**
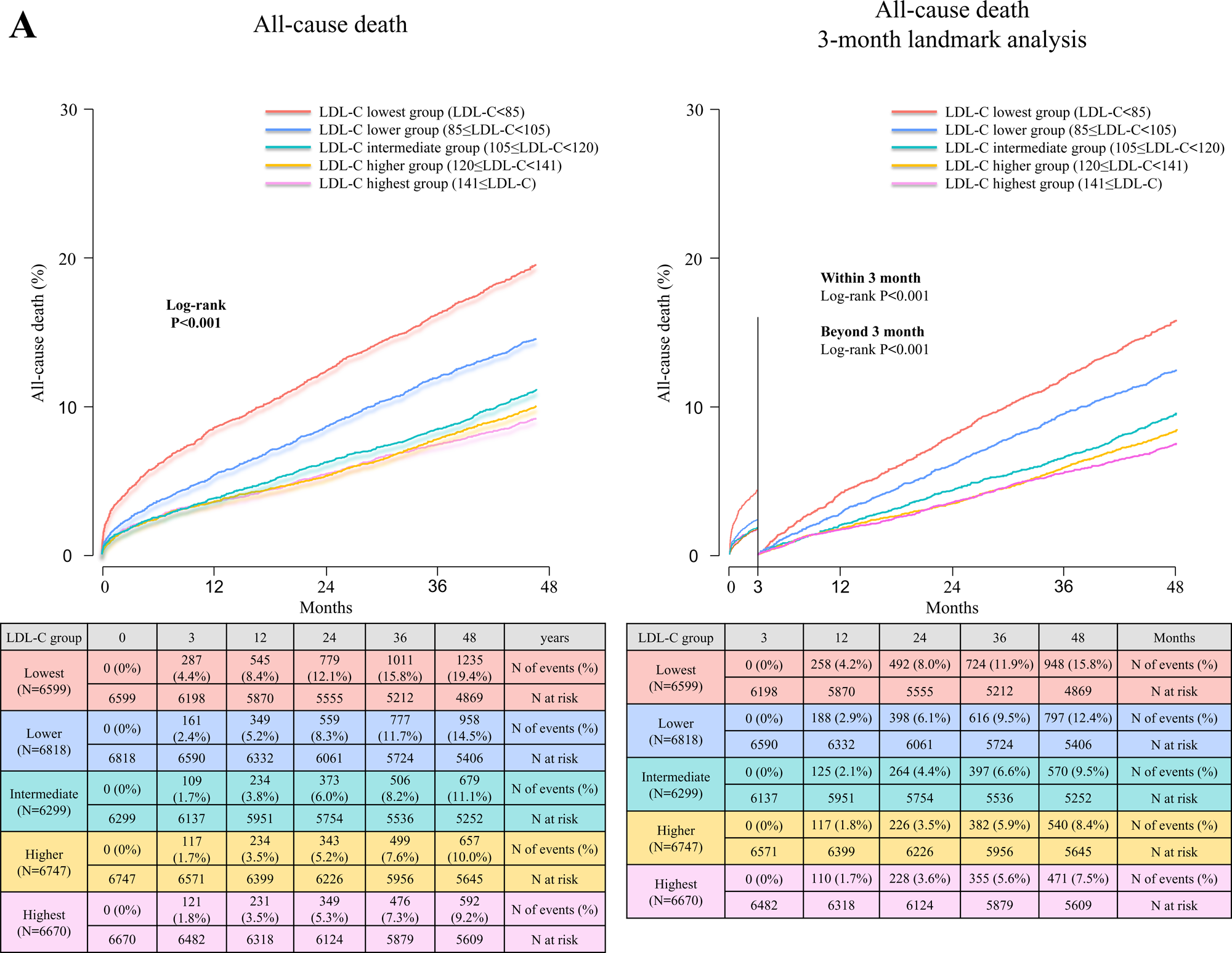
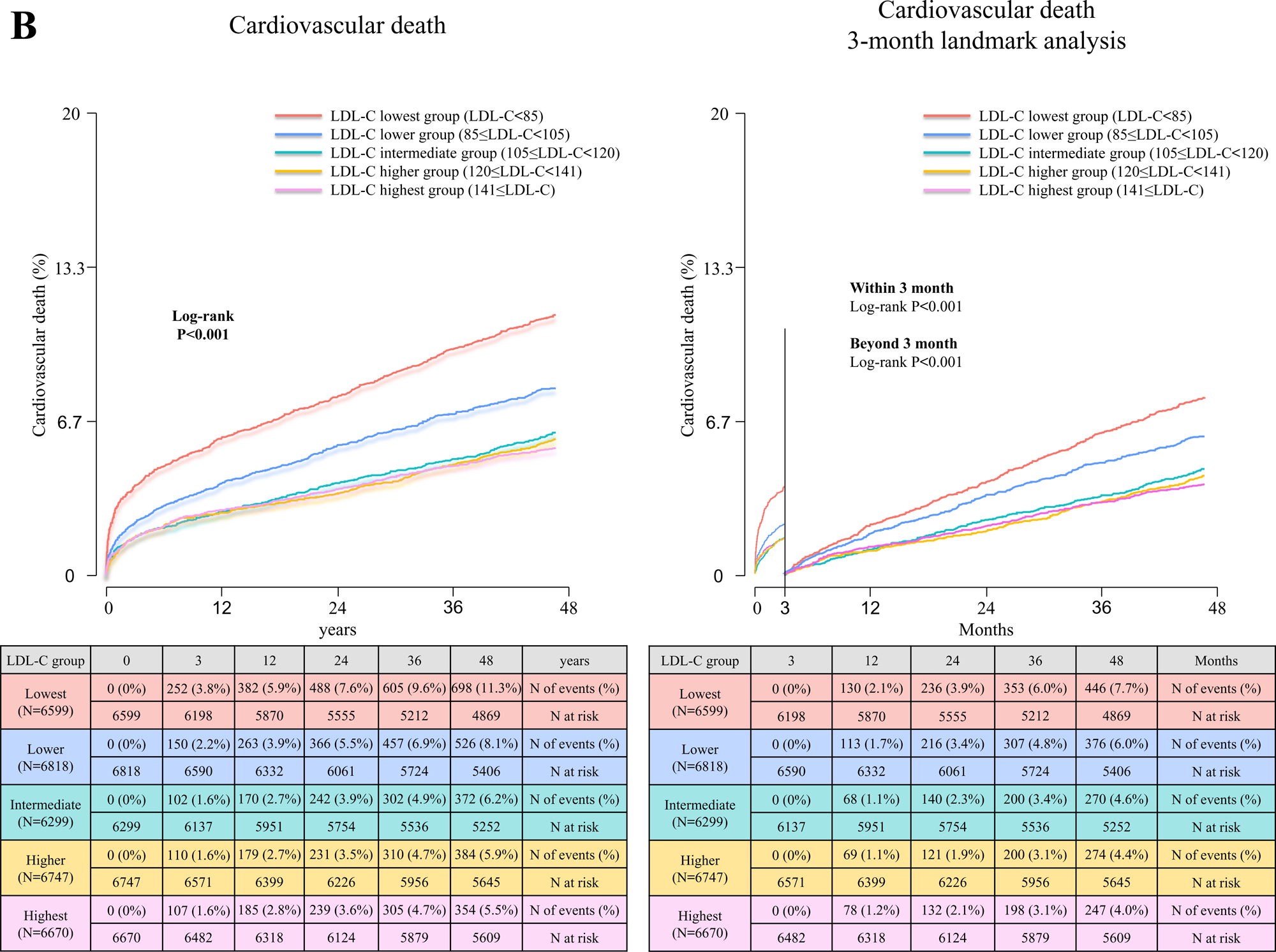
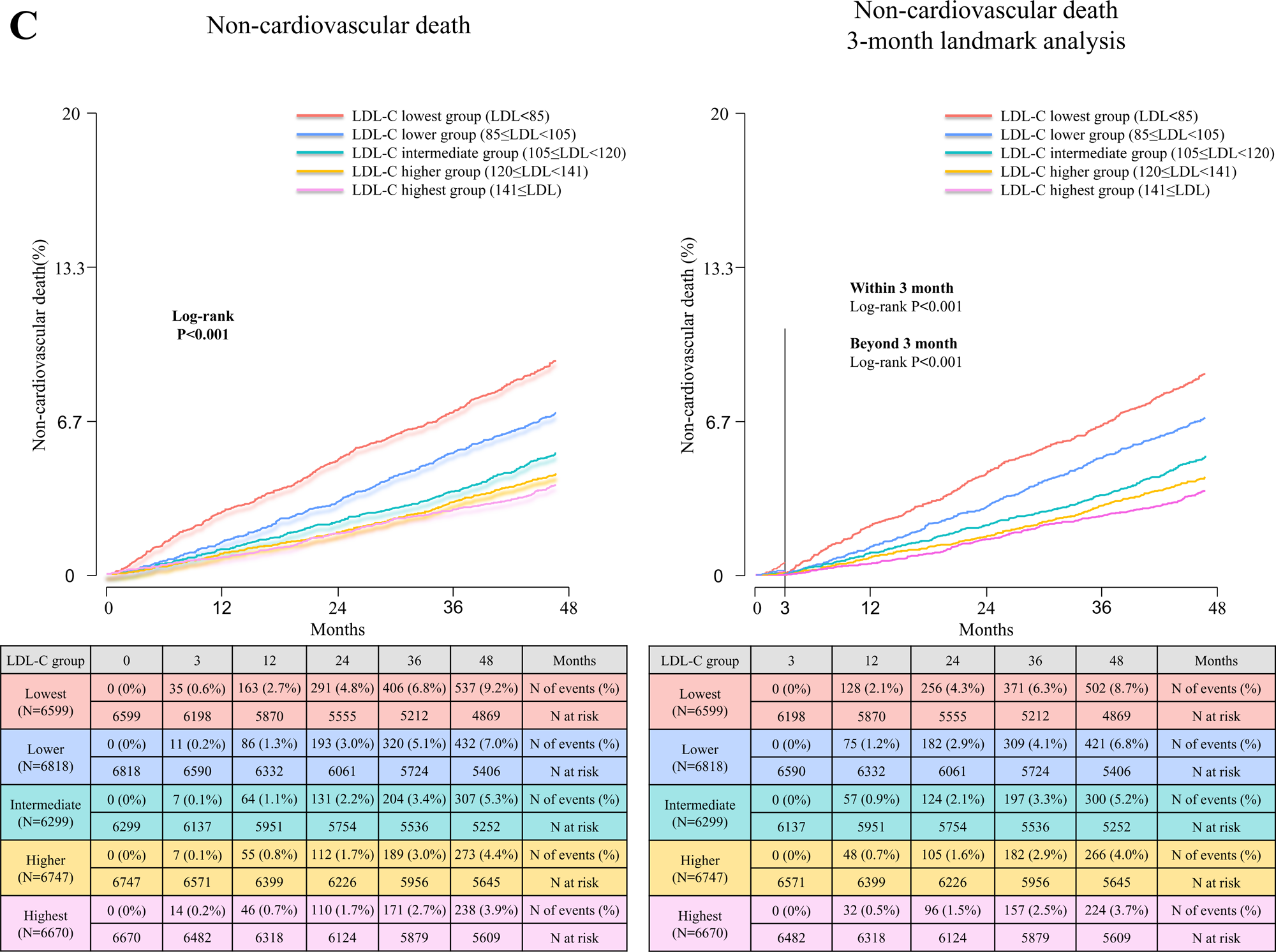
Kaplan–Meier curves for (A) all-cause death, (B) cardiovascular death and (C) non-cardiovascular death according to the LDL-C groups. Follow-up was censored at 4 years. The right-hand panels indicated the results from the 3-month landmark analyses. LDL-C=low-density lipoprotein cholesterol

**Figure 3.**
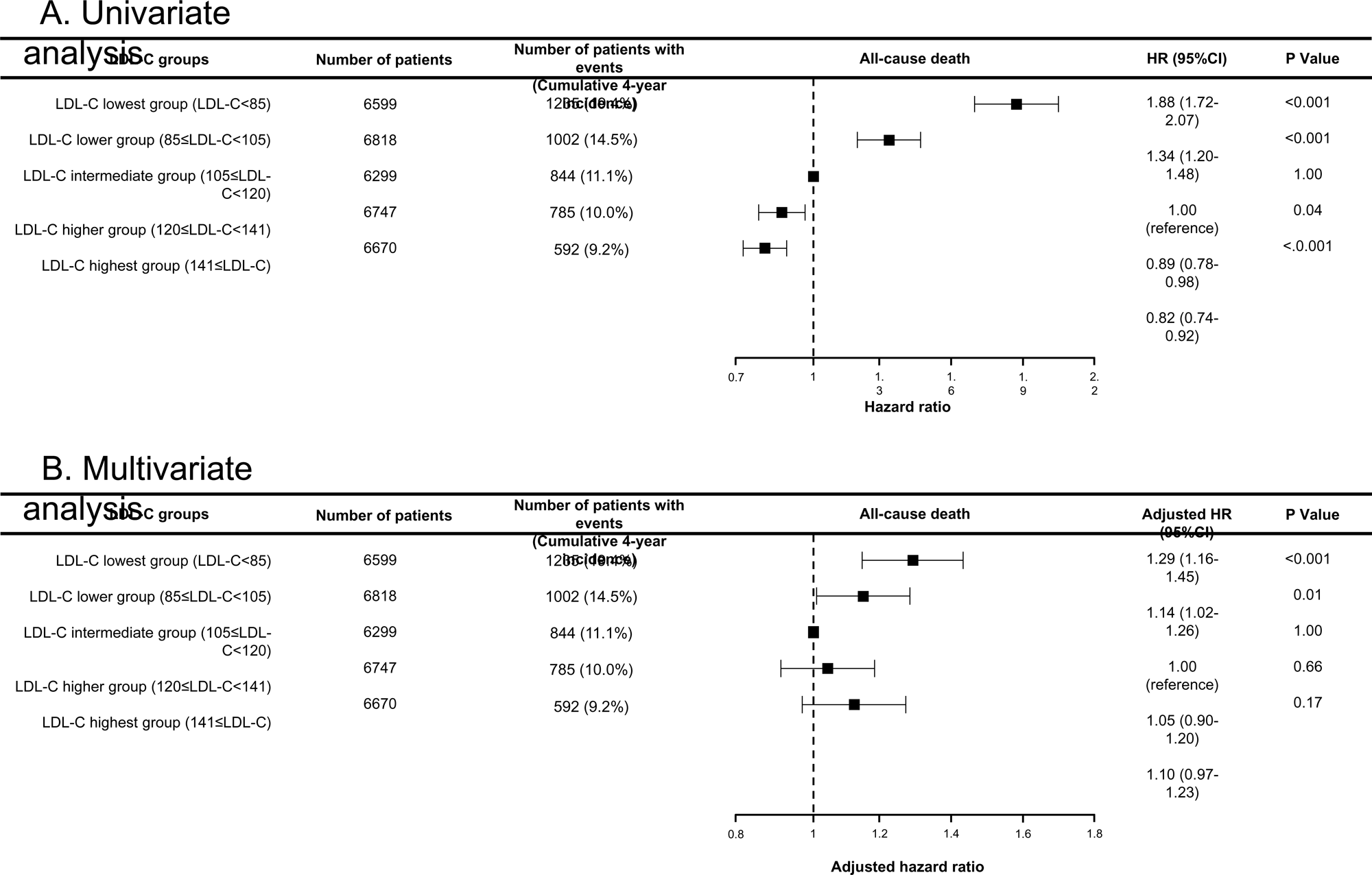
The risk of all-cause death according to the LDL-C groups (A) univariate cox regression models, and (B) multivariable cox regression models. The risk-adjusting factors in the multivariable variables were age, sex, body mass index (BMI), AMI presentation, smoker, hypertension, diabetes, acute heart failure, cardiogenic shock, ejection fraction, prior peripheral artery disease, prior heart failure, prior stroke, atrial fibrillation, chronic obstruction pulmonary disease, liver cirrhosis, malignancy, estimated glomerular filtration rate (eGFR), hemoglobin, platelet count, study cohorts, use of β blocker, use of renin-angiotensin system inhibitors, and use of any medications for dyslipidemia at hospitalization. LDL-C=low-density lipoprotein cholesterol, HR=hazard ratio, CI=confidence interval

**Table 2.** Clinical outcomes at 4-year according to the baseline LDL-C levels.

|  | Overall<br>(N=33133) | Baseline LDL-C (mg/dl) |  |  |  |  | Log-rank<br>P values | Log-rank trend<br>P values |
| --- | --- | --- | --- | --- | --- | --- | --- | --- |
|  |  | Quintiles |  |  |  |  |  |  |
|  |  | Lowest<br>group<br>(LDL-C <85) | Lower<br>group<br>(85≤ LDL-C <105) | Intermediate<br>group<br>(105≤ LDL-C <120) | Higher<br>group<br>(120≤ LDL-C <141) | Highest<br>group<br>(141≤ LDL-C) |  |  |
|  |  | N=6599 | N=6818 | N=6299 | N=6747 | N=6670 |  |  |
| Clinical outcomes |  |  |  |  |  |  |  |  |
| All-cause death | 4121 (12.8%) | 1235 (19.4%) | 1002 (14.5%) | 844 (11.1%) | 785 (10.0%) | 592 (9.2%) | <0.001 | <0.001 |
| Cardiovascular death | 2334 (7.4%) | 698 (11.3%) | 526 (8.1%) | 372 (6.2%) | 384 (5.9%) | 354 (5.5%) | <0.001 | <0.001 |
| Non-cardiovascular death | 1787 (5.9%) | 537 (9.2%) | 432 (7.0%) | 307 (5.3%) | 273 (4.4%) | 238 (3.9%) | <0.001 | <0.001 |
| Cardiac death | 1938 (6.1%) | 593 (9.6%) | 440 (6.8%) | 294 (4.9%) | 306 (4.7%) | 305 (4.8%) | <0.001 | <0.001 |
| Sudden death | 466 (1.5%) | 115 (2.0%) | 103 (1.7%) | 61 (1.1%) | 88 (1.4%) | 99 (1.6%) | <0.001 | <0.001 |
| Acute myocardial infarction | 1568 (5.0%) | 304 (5.1%) | 309 (4.8%) | 303 (5.0%) | 339 (5.3%) | 313 (4.9%) | 0.85 | 0.67 |
| Stroke | 1683 (5.5%) | 355 (6.0%) | 368 (5.9%) | 337 (5.8%) | 318 (5.0%) | 305 (5.0%) | 0.02 | 0.01 |
| Heart failure admission | 2045 (6.7%) | 503 (9.7%) | 448 (7.2%) | 365 (6.2%) | 352 (5.6%) | 377 (6.0%) | <0.001 | <0.001 |

Data are presented as number of patients with event (cumulative 4-year incidence).
3 LDL-C=low-density lipoprotein cholesterol.

**Table 3.**

| a | Univariate Cox regression analysis |  |  |  |  |  |  |  |  |
| --- | --- | --- | --- | --- | --- | --- | --- | --- | --- |
|  | Lowest group<br>(LDL-C <85)<br>N=6599 |  | Lower group<br>(85≤ LDL-C <105)<br>N=6818 |  | Intermediate group<br>(105≤ LDL-C <120)<br>N=6299 | Higher group<br>(120≤ LDL-C <141)<br>N=6747 |  | Highest group<br>(141≤ LDL-C)<br>N=6670 |  |
|  | HR (95% CI) | P values | HR (95% CI) | P values | HR (95% CI) | HR (95% CI) | P values | HR (95% CI) | P values |
| All-cause death | 1.86 (1.70-2.05) | <0.001 | 1.34 (1.21-1.48) | <0.001 | 1.00 (Reference) | 0.90 (0.80-1.00) | 0.054 | 0.82 (0.74-0.92) | <0.001 |
| Cardiovascular death | 1.90 (1.68-2.16) | <0.001 | 1.33 (1.17-1.52) | <0.001 | 1.00 (Reference) | 0.96 (0.83-1.11) | 0.58 | 0.90 (0.78-1.04) | 0.14 |
| Non-cardiovascular death | 1.82 (1.56-2.09) | <0.001 | 1.34 (1.16-1.55) | <0.001 | 1.00 (Reference) | 0.83 (0.70-0.97) | 0.02 | 0.73 (0.62-0.86) | <0.001 |
| Cardiac death | 2.04 (1.77-2.34) | <0.001 | 1.41 (1.22-1.63) | <0.001 | 1.00 (Reference) | 0.97 (0.83-1.15) | 0.70 | 0.98 (0.83-1.14) | 0.79 |
| Sudden death | 1.93 (1.42-2.64) | <0.001 | 1.60 (1.17-2.20) | 0.004 | 1.00 (Reference) | 1.34 (0.97-1.86) | 0.08 | 1.53 (1.11-2.10) | 0.009 |
| Acute myocardial infarction | 1.00 (0.85-1.17) | 0.97 | 0.96 (0.82-1.12) | 0.58 | 1.00 (Reference) | 1.04 (0.89-1.22) | 0.59 | 0.97 (0.83-1.14) | 0.74 |
| Stroke | 1.05 (0.91-1.22) | 0.51 | 1.02 (0.88-1.19) | 0.76 | 1.00 (Reference) | 0.88 (0.75-1.02) | 0.10 | 0.85 (0.73-0.99) | 0.04 |
| Heart failure admission | 1.42 (1.24-1.62) | <0.001 | 1.16 (1.01-1.34) | 0.03 | 1.00 (Reference) | 0.89 (0.77-1.04) | 0.13 | 0.97 (0.84-1.12) | 0.70 |
|  | Multivariate Cox regression analysis |  |  |  |  |  |  |  |  |
|  | Lowest group<br>(LDL-C <85)<br>N=6599 |  | Lower group<br>(85≤ LDL-C <105)<br>N=6818 |  | Intermediate group<br>(105≤ LDL-C <120)<br>N=6299 | Higher group<br>(120≤ LDL-C <141)<br>N=6747 |  | Highest group<br>(141≤ LDL-C)<br>N=6670 |  |

| Clinical outcomes | Adjusted HR<br>(95% CI) | P values | Adjusted HR<br>(95% CI) | P values | Adjusted HR<br>(95% CI) | Adjusted HR<br>(95% CI) | P values | Adjusted HR<br>(95% CI) | P values |
| --- | --- | --- | --- | --- | --- | --- | --- | --- | --- |
| All-cause death | 1.29 (1.16-1.44) | <0.001 | 1.15 (1.03-1.29) | 0.01 | 1.00 (Reference) | 1.10 (0.93-1.19) | 0.45 | 1.05 (0.97-1.25) | 0.15 |
| Cardiovascular death | 1.17 (1.01-1.35) | 0.04 | 1.09 (0.94-1.27) | 0.26 | 1.00 (Reference) | 1.04 (0.89-1.24) | 0.58 | 1.11 (0.94-1.32) | 0.23 |
| Non-cardiovascular death | 1.35 (1.15-1.60) | <0.001 | 1.19 (1.01-1.40) | 0.04 | 1.00 (Reference) | 0.99 (0.82-1.20) | 0.94 | 1.13 (0.93-1.37) | 0.23 |
| Cardiac death | 1.22 (1.04-1.43) | 0.01 | 1.13 (0.96-1.34) | 0.15 | 1.00 (Reference) | 1.03 (0.86-1.25) | 0.70 | 1.16 (0.96-1.40) | 0.13 |
| Sudden death | 1.44 (1.01-2.06) | 0.049 | 1.60 (1.10-2.32) | 0.01 | 1.00 (Reference) | 1.63 (1.10-2.41) | 0.02 | 1.90 (1.27-2.84) | 0.002 |
| Acute myocardial infarction | 0.92 (0.77-1.09) | 0.33 | 0.96 (0.81-) | 0.67 | 1.00 (Reference) | 1.16 (0.97-1.38) | 0.10 | 1.11 (0.93-1.33) | 0.26 |
| Stroke | 0.95 (0.80-1.13) | 0.57 | 1.01 (0.85-1.20) | 0.91 | 1.00 (Reference) | 0.98 (0.81-1.17) | 0.81 | 0.94 (0.78-1.13) | 0.53 |
| Heart failure admission | 1.11 (1.01-1.22) | 0.04 | 1.07 (0.92-1.25) | 0.35 | 1.00 (Reference) | 0.98 (0.83-1.16) | 0.80 | 1.31 (1.12-1.54) | 0.001 |

<sup>1</sup> HR=hazard ratio, CI=confidence interval

The results of the unadjusted penalized smoothing splines showed that the risk of all-cause death was greatly increased by a decrease in LDL rather than an increase in LDL (**Supplemental Figure 2**). The adjusted penalized smoothing splines showed a U-shaped association between continuous LDL-C levels and the risk of all-cause death (**Figure 4A**). Similarly, a U-shaped association was observed between cardiovascular and non-cardiovascular deaths (**Figure 4B and 4C**). The excess adjusted risk of extremely high LDL-C levels (> 200 mg/dL) appeared to be high for all-cause, cardiovascular, and non-cardiovascular mortality.

**Figure 4.**
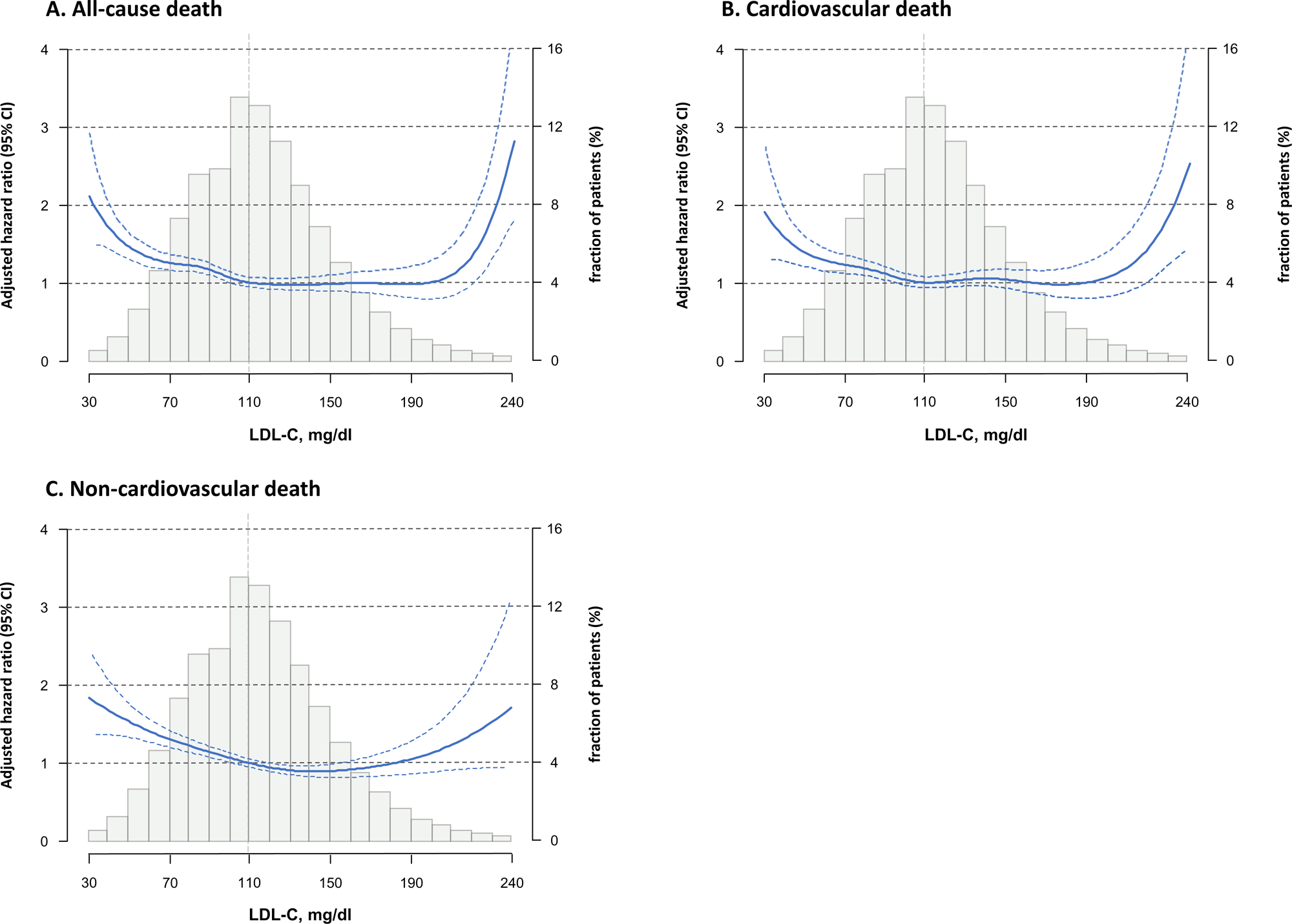
Penalized spline curves on a continuous scale for the adjusted hazard ratios for (A) all-cause death, (B) cardiovascular death, (C) non-cardiovascular death. The adjusted factors were age, sex, body mass index (BMI), AMI presentation, smoker, hypertension, diabetes, acute heart failure, cardiogenic shock, ejection fraction, prior peripheral artery disease, prior heart failure, prior stroke, atrial fibrillation, chronic obstruction pulmonary disease, liver cirrhosis, malignancy, estimated glomerular filtration rate (eGFR), hemoglobin, platelet count, study cohorts, use of β blocker, use of renin-angiotensin system inhibitors, and use of any medications for dyslipidemia at hospitalization. The histograms show the fraction of patients with different LDL levels.

These results were consistent in a sensitivity analysis that divided the study patients into seven groups according to the percentiles of the distribution of LDL-C levels, the 5^th^, 20^th^, 40^th^, 60^th^, 80^th^, and 95^th^ percentiles (**Supplemental Figure 3 and 4)**. In the subgroup analyses, no significant interactions were observed regarding lipid-lowering treatment during hospitalization (P for interaction=0.08), age (P for interaction=0.08), sex (P for interaction=0.91), AMI presentation (P for interaction=0.36), or study cohorts (P for interaction=0.87) (**Supplemental Figure 5**). Nevertheless, the magnitude of the excess mortality risk in the lowest LDL-C group relative to the intermediate LDL-C group was greater in patients not receiving lipid-lowering therapy than in those receiving lipid-lowering therapy at index hospitalization.

The PDP, based on gradient boosting for all-cause death, revealed no significant interaction between LDL-C levels and other risk factors such as age, BMI, HbA1c levels, systolic blood pressure, eGFR, and hemoglobin levels (**Figure 5**). The marginal effect of the two variables of interest on all-cause death based on PDP showed that approximate normal and appropriate values were associated with the lowest risk, and outliers were associated with high risk.

**Figure 5.**
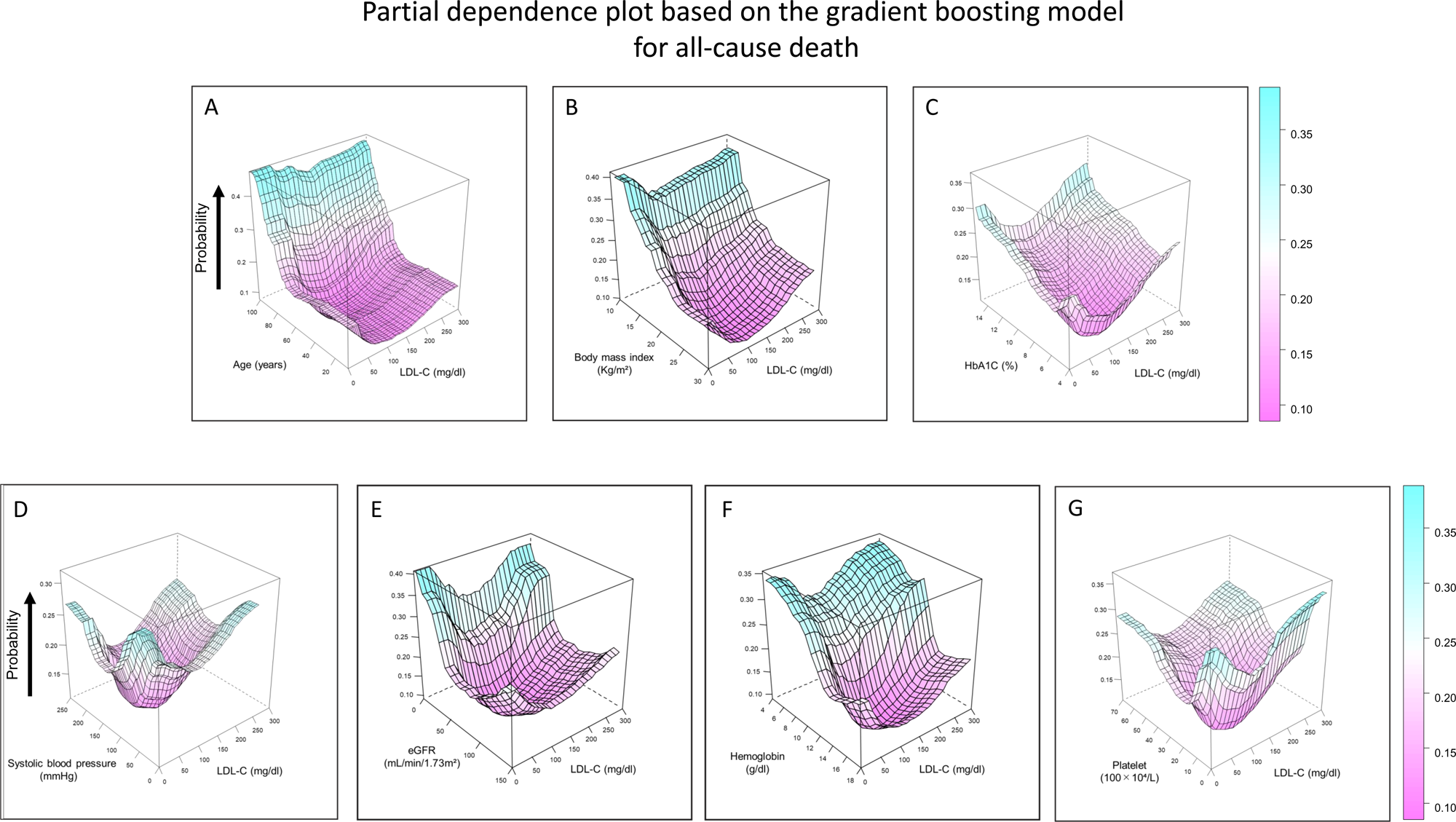
Partial dependence plot for all-cause death to visualized interaction between LDL-C and (A) age, (B) body mass index, (C) HbA1c, (D) systolic blood pressure, (E) eGFR, (F) hemoglobin, and (G) platelet according to gradient boosting. Partial dependence plot based on gradient boosting model, which shows the marginal effect of two predictors on the prediction, was used to visualize the association and interaction between LDL-C and the other predictors for all-cause death. The adjusted factors were age, sex, body mass index (BMI), AMI presentation, smoker, hypertension, diabetes, acute heart failure, cardiogenic shock, ejection fraction, prior peripheral artery disease, prior heart failure, prior stroke, atrial fibrillation, chronic obstruction pulmonary disease, liver cirrhosis, malignancy, estimated glomerular filtration rate (eGFR), hemoglobin, platelet count, cohort number, use of β blocker, use of renin-angiotensin system inhibitors, and use of any medications for dyslipidemia at hospitalization. LDL-C=low-density lipoprotein cholesterol.

## Discussion

The present study showed that among those who underwent coronary revascularization, patients with lower LDL-C levels had more comorbidities and a significantly higher risk of all-cause death, irrespective of the mode of death. These findings are in line with those of previous studies showing the association of low LDL-C levels with clinical conditions with poor prognosis, such as cardiogenic shock and malignancy.^24–28^ The Lipid Paradox, which indicates a high risk of mortality with low LDL-C levels, has been reported in several studies. A study from the National Registry of Myocardial Infarction involving 115 492 patients showed that the risk of in-hospital mortality in the second to fourth quartiles was lower than that in the lowest quartile of LDL-C (<77 mg/dl).^29^ A study from the Korean registry of acute myocardial infarction involving 5532 patients also showed that a baseline LDL-C level of <70 mg/dL was significantly associated with a high incidence of cardiovascular events after AMI.^30^ Other short-term or small-scale studies focusing on LDL-C levels and mortality have been conducted; however, real-world data are limited to low baseline LDL-C levels and long-term clinical outcomes. To the best of our knowledge, this is the first study to demonstrate the association between low LDL-C levels and a higher adjusted risk of various clinical events, including all-cause death, cardiovascular death, non-cardiovascular death, admission for heart failure, and sudden death in patients with coronary artery disease. The present study also showed that the higher risk of death in patients with low LDL-C levels was driven by both early and late events. Furthermore, we closely examined the baseline characteristics of patients with low LDL-C levels and their association with mortality, including the interactions between LDL-C and other variables.

Severe comorbidities in patients with lower LDL-C levels may partially explain their higher risk of all-cause death. However, the higher risk of all-cause death among patients with lower LDL-C levels persisted even after adjusting for baseline characteristics. Moreover, 3-month landmark analyses further confirmed the higher risk for all-cause death beyond 3 months in the very low LDL-C quintile, indicating that acute high-risk clinical characteristics, such as the presence of cardiogenic shock, were not the main drivers for the higher risk of all-cause death in patients with lower LDL-C levels. Notably, there were no significant interactions between various demographic and clinical factors, suggesting that low LDL-C levels are an independent risk factor for all-cause death across a broad range of patient populations.

The mechanisms underlying poor clinical outcomes in patients with low LDL-C levels are not yet fully understood. There are several possible explanations for the higher mortality risk observed in these patients, including frailty, malignancy, and chronic inflammation. Frailty is known to be associated with low LDL-C levels, which might explain the higher risk of admission for heart failure and subsequent cardiovascular mortalities.^31, 32^ The strong relationship between low LDL-C levels and malignancy might also explain the higher non-cardiovascular mortality in patients with low LDL-C levels.^16^ Since the risk of malignancy was higher than that of the general population due to overlapping risk factors for atherosclerosis and malignancy,^33, 34^ systemic examination focusing on malignancy in patients with low LDL-C levels might be important in mitigating the non-cardiovascular death risks after PCI. Chronic inflammation is also strongly associated with low LDL-C levels,^26, 30, 35^ and has been reported to be a strong predictor of cardiovascular events among those who have already received contemporary statins.^36, 37^

Although there is ample evidence supporting the benefit of lipid-lowering therapy using the “lower the better” LDL-C approach in high-risk patients, the efficacy of this approach in patients with very low baseline LDL-C levels remains unclear.^8, 38, 39^ Current clinical guidelines recommend adding non-statin LDL-C lowering agents to statin therapy in very high-risk patients when the LDL-C levels remain at 70 mg/dL or above. Consequently, patients with low baseline LDL-C levels are less likely to be prescribed aggressive lipid-lowering therapy despite their higher risk of all-cause death. It is worth noting that patients with low baseline LDL-C levels are presumably less likely to be enrolled in randomized controlled trials due to their poorer conditions and prognosis.^40^ Our all-comer observational study highlighted that lower baseline LDL-C levels were associated with more comorbidities and a significantly higher risk of all-cause death in patients undergoing coronary revascularization, indicating that there was potential room for improvement in their management. Further randomized controlled trials are warranted to elucidate the need for preemptive interventions, such as aggressive lipid-lowering therapy or anti-inflammatory medications in this particular population.

### Limitations

This study has several limitations that should be considered when interpreting the results. First, the lack of available LDL-C levels in approximately 15% of the patients introduced the possibility of selection bias, as this subgroup may differ from those with available LDL-C data. Second, caution is required in the interpretation of the results obtained through penalized splines because patients at both ends of the spline curve were scarce. Third, there might remain a possibility of unadjusted confounders that could influence the observed associations. These confounders might include not only clinical conditions but also other factors, such as timing and method of measurement, as well as dietary status. Fourth, high-intensity statin therapy was rarely implemented in our study population. The observations from our study might be changed by the effect of high-intensity statin therapy with or without additional aggressive LDL-C lowering therapy, which is recommended in the current guidelines. Finally, LDL-C levels are influenced by many factors, including the timing and method of measurement, lipid-lowering treatments, and diet, and these factors were not rigorously adjusted for in this study. A substantial proportion of patients received lipid-lowering therapy at the time of index hospitalization, which might have affected the characteristics of patients with lower LDL-C levels. Nevertheless, the magnitude of the excess mortality risk in the lowest LDL-C group relative to the intermediate LDL-C group was greater in patients not receiving lipid-lowering therapy than in those receiving lipid-lowering therapy at index hospitalization.

## Conclusions

Lower baseline LDL-C levels were associated with more comorbidities and a significantly higher risk of death, regardless of cardiovascular or non-cardiovascular causes, in patients who underwent coronary revascularization.

## Data Availability

The deidentified participant data will not be shared.

## Acknowledgments

The authors thank the clinical research coordinators in the Research Institute for Production Development.

## Abbreviations

ACS: Acute coronary syndrome
AMI: Acute myocardial infarction
BMI: Body mass index
CABG: Coronary artery bypass grafting
CREDO-Kyoto: Coronary Revascularization Demonstrating Outcome Study in Kyoto
LDL-C: Low density lipoprotein cholesterol
PCI: Percutaneous coronary intervention PDP = Partial dependence plot

